# Healthcare workers preparedness for COVID-19 pandemic in the occupied Palestinian territory: a cross-sectional survey

**DOI:** 10.1101/2020.05.09.20096099

**Authors:** Osaid Alser, Heba Alghoul, Zahra Alkhateeb, Ayah Hamdan, Loai Albarqouni, Kiran Saini

## Abstract

**Background:** The coronavirus disease 19 (COVID-19) pandemic threatens to overwhelm the capacity of the vulnerable healthcare system in the occupied Palestinian territory (oPt). Sufficient training of healthcare workers (HCWs) in how to manage COVID-19 and the provision of personal protective equipment (PPE) to enable them to do so will be key tools in allowing oPt to mount a credible response to the crisis.

**Methods:** A cross-sectional study was conducted using a validated online questionnaire. Data collection occurred between March 30, 2020 and April 12, 2020. The primary outcomes were availability of PPE and HCWs preparedness in oPt for COVID-19 pandemic. The secondary outcome was the regional and hospital differences in oPt in terms of availability of PPE and HCWs preparedness.

**Results:** Of 138 respondents, only 38 HCWs (27.5%) always had access to facemasks when needed and 15 (10.9%) for isolation gowns. The vast majority of HCWs did not find eye protection (n=128, 92.8%), N95 respirators (n=132, 95.7%), and face shields (n=127, 92%) always available. Compared to HCWs in West Bank, those in the Gaza Strip were significantly less likely to have access to alcohol sanitizers (p=0.026) and gloves (p <0.001). On average, governmental hospitals were significantly less likely to have all appropriate PPE measures than non-governmental institutions (p = 0.001). As for preparedness, only 16 (11.6%) surveyed felt confident in dealing with a potential COVID-19 case. With 57 (41.3%) having received any COVID-19 related training and 57 (41.3%) not having a local hospital protocol.

**Conclusion:** HCWs in oPt are underprepared and severely lacking adequate PPE provision. The lack of local protocols, and training has left HCWs confidence exceedingly low. The lack of PPE provision will exacerbate spread of COVID-19 and deepen the crisis, whilst putting HCWs at risk.

## Background

With the ongoing coronavirus disease 2019 (COVID-19) pandemic, the humanitarian and healthcare crisis in low-to-middle income countries (LMICs) such as the occupied Palestinian territory (oPt) is expected to be amplified and this will further cripple the healthcare system. As of May 7, 2020, the World Health Organization (WHO) has recorded 547 confirmed cases of COVID-19 in the oPt; 527 in the West Bank and 20 in the Gaza Strip – with 4 fatalities.^1^

The United Nations Relief and Works Agency (UNRWA) has been unable to support Palestinians’ COVID-19 response needs at their full capacity at the consequence of funding cut and legal restrictions that were in place prior to the pandemic.^2^ Multiple COVID-19 testing sites serving Palestinians in East Jerusalem have been closed by the Israeli authorities.^3^ The West Bank is particularly vulnerable due to checkpoint closures, halt of the transportation of patients to hospitals, and redistribution of clinical supplies. The Gaza Strip is one of the most densely populated places on earth with 2 million inhabitants, mostly refugees, live in 365 sq. km^2^, allowing for an accelerated spread of disease should a COVID-19 outbreak manifest.^4^ Other LMICs in the Middle East and Africa have also reported scarcity of personal protective equipment (PPE) for front line healthcare workers (HCWs).^5, 6^

We hypothesize that (HCWs) in the oPt are largely underprepared to address COVID-19 related needs of the Palestinian population in both the West Bank and Gaza Strip. Shortages of PPE pose a serious threat to COVID-19 containment in the oPt. It is also expected that HCWs in the oPt have likely received insufficient training on how to address spread and containment of COVID-19; institutions themselves may not have yet been equipped to draw up or implement preventative or management protocols. To the best of our knowledge, there have been no studies evaluating the preparedness of the HCWs in the oPt to face COVID-19 pandemic. In this study, we aim to evaluate the availability of PPE and the level of preparedness among the HCWs in the oPt.

## Methods

### Study design, setting and data collection

We conducted a cross-sectional study using an online survey tool. Our survey (*Additional material - Table 1*) was modified from two validated questionnaires; the first was utilized during the H1N1 influenza pandemic ^7^ and the second one was the Personnel, Infrastructure, Procedures, Equipment and Supplies (PIPES) surgical capacity assessment tool. ^8^ Our modified questionnaire consisted of 22 questions divided into 3 different sections (respondent and healthcare facility characteristics, availability of PPE and HCWs preparedness). Availability of PPE and HCWs preparedness were assessed on a 5-point Likert scale. The questionnaire was distributed to HCWs in the oPt through convenient sampling between March 30, 2020 and April 12, 2020. E-mail lists for participants in an educational link (OxPal) and social media (Facebook, Twitter, and LinkedIn) groups of HCWs in oPt were used to disseminate the questionnaire. Participants were required to sign in to limit the number of responses to one per respondent.

**Table 1.** Healthcare workers and their healthcare facility characteristics

| Parameter | Value |
| --- | --- |
| **Age, median (IQR) - [range]** | 28 (24, 35) - [19-57] |
| **Male, n (%)** | 85 (61.6) |
| **Regional Location, n (%)** |  |
| *Gaza Strip* | 97 (70.3) |
| *West Bank (including East Jerusalem)* | 41 (29.7) |
| **Profession, n (%)** |  |
| *Medicine* | 69 (50) |
| *Nursing* | 35 (25.4) |
| *Physiotherapy* | 13 (9.4) |
| *Dentistry* | 6 (4.3) |
| *Other\** | 15 (10.9) |
| **Level of training, n (%)** |  |
| *Student* | 23 (16.7) |
| *Trainee* | 23 (16.7) |
| *Non-specialized* | 69 (50.0) |
| *Specialized* | 23 (16.7) |
| **Department / Specialty, n (%)** |  |
| *Emergency medicine* | 20 (14.5) |
| *Surgery (including sub-specialties)* | 19 (13.8) |
| *Family medicine (primary care)* | 14 (10.1) |
| *Internal medicine (including sub-specialties)* | 13 (9.4) |
| *Pediatrics* | 10 (7.2) |
| *Dentistry* | 8 (5.8) |
| *Anesthesiology/critical care* | 7 (5.1) |
| *Obstetrics/Gynecology* | 7 (5.1) |
| *Radiology* | 6 (4.3) |
| *Other\** | 13 (9.4) |
| *Not applicable (e.g. students)* | 21 (15.2) |
| **Type of healthcare facility (I), n (%)** |  |
| *Primary healthcare center/ clinic* | 43 (31.2) |
| *Secondary hospital* | 29 (21) |
| *Tertiary (referral) hospital* | 63 (45.7) |
| *Isolation Center* | 1 (0.7) |
| *Not applicable* | 2 (1.4) |
| **Type of healthcare facility (II), n (%)** |  |
| *Governmental (Ministry of Health)* | 98 (71) |
| *Private* | 26 (18.8) |
| *Non-governmental (NGO) or Mission* | 13 (9.4) |
| *Not applicable* | 1 (0.7) |

\*Includes: Optometry, medical secretary, medical laboratory, pharmacy, radiography.

### Study outcomes

The primary outcomes assessed were availability of PPE and HCWs preparedness in oPt in the era of COVID-19 pandemic. The secondary outcome was to assess the differences between Gaza Strip and West Bank, and between governmental and non-governmental in oPt in terms of availability of PPE and HCWs preparedness to face the COVID-19 pandemic.

### Statistical analysis

Respondent characteristics were summarized using descriptive statistics. For continuous data, mean and standard deviation (SD) were used to report normally distributed data, while median and interquartile ranges (IQR) were used for non-normally distributed data. For categorical data, results were summarized as counts (n) and percentages (cumulative incidence). Univariate analysis (chi-squared and Fisher’s Exact tests) was also used to compare participants’ profession, geographical location, and responses to questions related to the availability of PPE and HCWs preparedness for the COVID-19 pandemic. Likert scale variables were converted from 5-point to binary variables for univariate analysis. For example, often available, sometimes available, rarely availably and never available were grouped together as ‘not always available’. Strongly agree and moderately agree were grouped into ‘agree’ variable compared to ‘neutral’ and ‘disagree’ categories. Missing data were considered missing completely at random, therefore we performed complete case analysis. All statistical analyses were performed using IBM Corp. Released 2019. IBM SPSS Statistics for Windows, Version 26.0. Armonk, NY: IBM Corp.

## Results

Of 140 completed surveys, two were excluded from the study as they were either working outside the oPt or in a non-medical profession.

### HCWs and their healthcare facility characteristics

Of 138 HCWs included in the study, 97 respondents (70.3%) were from Gaza Strip and 41 (29.7%) were from the West Bank. The median (IQR) age was 28 (24-35) years with a range from 19 to 57 years old. 85 respondents (61.6 %) were males. Exactly half of respondents were medical doctors, with approximately 35 (25.4%) in nursing and the remaining quarter in physiotherapy, dentistry, or another health-related profession. 20 (14.5%) of the respondents worked in emergency medicine and 19 (13.8%) in surgery, 14 (10.1%) in primary care and 13 (9.4%) in internal medicine. With regards to place of work, 63 (45.7%) of the respondents worked in a tertiary hospital, 29 (21%) in a secondary facility and 43 (31%) in a primary healthcare center or clinic. One respondent worked in a COVID-19 isolation center. 98 (71%) worked in a governmental institution operated by the Ministry of Health, 26 (18.8%) worked in a private hospital and 13 (9.4%) in a non-governmental organization (NGO) or mission-based place of care (Table 1).

### Availability of PPE and HCWs preparedness in terms of infection control training

Only 67 (48.6%) and 71 (51.4%) of HCWs surveyed indicated that they always had alcohol-based sanitizer and gloves available in their institutions, respectively. Only 38 (27.5%) of respondents indicated that regular face masks were always available when needed, and just over 15 (10.9%) of respondents reported that isolation gowns were always available in their institutions. Over 128 (92.8%), 132 (95.7%), and 127 (92%) of respondents indicated that eye protection, N95 respirators, and face shields were not always available to them at their institutions, respectively. 57 (41.3%) of HCWs surveyed indicated that their hospital did not provide a local protocol for the management of COVID-19. Only 57 (41.3%) of respondents had received any COVID-19 related training courses by the time of survey administration. 16 (11.6%) of HCWs surveyed agreed with the statement of feeling confident or well-prepared to deal with a potential COVID-19 case (Table 2).

**Table 2.** Availability of PPE and preparedness of healthcare workers in oPt for COVID-19 pandemic

| **Parameter** | **n (%)** |
| --- | --- |
| **Alcohol sanitizer** |  |
| *Always available* | 67 (48.6) |
| *Not always available* | 71 (51.4) |
| **Gloves** |  |
| *Always available* | 71 (51.4) |
| *Not always available* | 67 (48.6) |
| **Facemask** |  |
| *Always available* | 38 (27.5) |
| *Not always available* | 100 (72.5) |
| **N95 respirator** |  |
| *Always available* | 6 (4.3) |
| *Not always available* | 132 (95.7) |
| **Isolation gowns** |  |
| *Always available* | 15 (10.9) |
| *Not always available* | 123 (89.1) |
| **Eye protection (goggles/glasses)** |  |
| *Always available* | 10 (7.2) |
| *Not always available* | 128 (92.8) |
| **Face shields** |  |
| *Always available* | 11 (8) |
| *Not always available* | 127 (92) |
| **All protective measures (above)** |  |
| *Always available* | 15 (10.9) |
| *Not always available* | 123 (89.1) |
| **Receipt of any COVID-19 training course (e.g. infection control)** |  |
| *Yes* | 57 (41.3) |
| *No* | 81 (58.7) |
| **Hospital has provided local protocol for the management of COVID-19** |  |
| *Yes* | 81 (58.7) |
| *No* | 57 (41.3) |
| **I feel confident / well prepared dealing with a potential COVID-19 case** |  |
| *Agree* | 16 (11.6) |
| *Neutral / Disagree* | 122 (88.4) |

### Univariate analysis comparing Gaza Strip and West Bank in terms of availability of PPE and HCWs preparedness in terms of infection control training

Compared to the West Bank, respondents from the Gaza Strip reported significantly greater lack of alcohol-based hand sanitizers (p=0.03) and gloves (p<0.001), but no meaningful differences were observed between regions on other PPE or infection control readiness (Table 3).

**Table 3.** Univariate analysis comparing Gaza Strip and West Bank in terms of healthcare workers preparedness and personal protective equipment (PPE) availability

| Parameter | Gaza Strip, n (%)<br>N=97 | West Bank, n (%)<br>N=41 | p-value |
| --- | --- | --- | --- |
| **Alcohol sanitizer** |  |  |  |
| *Always available* | 41 (42.3) | 26 (63.4) | 0.026 |
| *Not always available* | 56 (57.7) | 15 (36.6) |  |
| **Gloves** |  |  |  |
| *Always available* | 40 (41.2) | 31 (75.6) | <0.001 |
| *Not always available* | 57 (58.8) | 10 (24.4) |  |
| **Facemask** |  |  |  |
| *Always available* | 24 (24.7) | 14 (34.1) | 0.299 |
| *Not always available* | 73 (75.3) | 27 (65.9) |  |
| **N95 respirator** |  |  |  |
| *Always available* | 5 (5.2) | 1 (2.4) | 0.669 |
| *Not always available* | 92 (94.8) | 40 (97.6) |  |
| **Isolation gowns** |  |  |  |
| *Always available* | 10 (10.3) | 87 (212.2) | 0.769 |
| *Not always available* | 5 (5.2) | 36 (87.8) |  |
| **Eye protection** |  |  |  |
| *Always available* | 8 (8.2) | 2 (4.9) | 0.723 |
| *Not always available* | 89 (91.8) | 39 (95.1) |  |
| **Face shields** |  |  |  |
| *Always available* | 10 (10.3) | 1 (2.4) | 0.174 |
| *Not always available* | 87 (89.7) | 40 (97.6) |  |
| **All PPE any other measures (above)** |  |  |  |
| *Always available* | 12 (12.4) | 3 (7.3) | 0.552 |
| *Not always available* | 85 (87.6) | 38 (92.7) |  |
| **Receipt of any COVID-19 training course (e.g. infection control)** |  |  |  |
| *Yes* | 44 (45.4) | 13 (31.7) | 0.185 |
| *No* | 53 (54.6) | 28 (68.3) |  |
| **Hospital has provided local protocol for the management of COVID-19** |  |  |  |
| *Yes* | 58 (59.8) | 23 (56.1) | 0.709 |
| *No* | 39 (40.2) | 18 (43.9) |  |
| **I feel confident / well prepared dealing with a potential COVID-19 case** |  |  |  |
| *Agree* | 13 (13.4) | 3 (7.3) | <0.001 |
| *Neutral/disagree* | 84 (86.6) | 38 (92.7) |  |

### Univariate analysis comparing governmental and non-governmental hospitals in terms of availability of PPE and HCWs preparedness in terms of infection control training

On average, governmental hospitals run by the MOH were also reported by respondents to be significantly lacking in sanitizer, gloves, facemasks, eye protection, and face shields compared to non-governmental institutions (p<0.05) (Table 4).

**Table 4.** Univariate analysis comparing Governmental and non-governmental hospitals in oPt in terms of healthcare workers preparedness and personal protective equipment (PPE) availability

| Parameter | Governmental hospitals, n (%)<br>N=98 | Non-governmental hospital, n (%)<br>N=39 | p-value |
| --- | --- | --- | --- |
| **Alcohol sanitizer** |  |  |  |
| *Always available* | 35 (35.7) | 32 (82.1) | <0.001 |
| *Not always available* | 63 (64.3) | 7 (17.9) |  |
| **Gloves** |  |  |  |
| *Always available* | 38 (38.8) | 32 (82.1) | <0.001 |
| *Not always available* | 60 (61.2) | 7 (17.9) |  |
| **Facemask** |  |  |  |
| *Always available* | 15 (15.3) | 23 (59.0) | <0.001 |
| *Not always available* | 83 (84.7) | 16 (41.0) |  |
| **N95 respirator** |  |  |  |
| *Always available* | 3 (3.1) | 3 (7.7) | 0.352 |
| *Not always available* | 95 (96.9) | 36 (92.3) |  |
| **Isolation gowns** |  |  |  |
| *Always available* | 8 (8.2) | 7 (17.9) | 0.129 |
| *Not always available* | 90 (91.8) | 32 (82.1) |  |
| **Eye protection** |  |  |  |
| *Always available* | 2 (2.0) | 8 (20.5) | 0.001 |
| *Not always available* | 96 (98.0) | 31 (79.5) |  |
| **Face shields** |  |  |  |
| *Always available* | 4 (4.1) | 7 (17.9) | 0.012 |
| *Not always available* | 94 (95.9) | 32 (82.1) |  |
| **All PPE any other measures (above)** |  |  |  |
| *Always available* | 5 (5.1) | 10 (25.6) | 0.001 |
| *Not always available* | 93 (94.9) | 29 (74.4) |  |
| **Receipt of any COVID-19 training course (e.g. infection control)** |  |  |  |
| *Yes* | 37 (37.8) | 19 (48.7) | 0.254 |
| *No* | 61 (62.2) | 20 (51.3) |  |
| **Hospital has provided local protocol for the management of COVID-19** |  |  |  |
| *Yes* | 57 (58.2) | 23 (59.0) | 1.00 |
| *No* | 41 (41.8) | 16 (41.0) |  |
| **I feel confident / well prepared dealing with a potential COVID-19 case** |  |  |  |
| *Agree* | 10 (10.2) | 6 (15.4) | 0.556 |
| *Neutral / Disagree* | 88 (89.8) | 33 (84.6) |  |

## Discussion

Our study demonstrates that the availability of PPE in both Gaza and the West Bank is insufficient to support the COVID-19 response needs of the oPt. Alcohol-based hand sanitizers, gloves, face masks, eye protection, isolation gowns, N95 respirators and face shields were reported to be inconsistently available, despite being internationally recommended as critical equipment needed for protecting health care workers from infection.^9^ Governmental hospitals, as opposed to non-governmental settings, appear to be particularly lacking in equipment. Lessons from prior outbreaks have underlined the importance of PPE in infection control.^10^ Recommendations from the WHO suggest the inadequate supply of infection prevention and control measures is vital to address immediately, with assistance from international partners if necessary.^11^ The WHO specifically mentioned supplies needed to implement recommended protocols, such as PPE, being a key resource to all national authorities currently not producing sufficient volumes themselves. Suggestions for other methods of procurement, conservation and management of PPE have been extensively covered in the literature during the pandemic.^12^ Many of these suggestions may not be viable in the geopolitical and economic context in which oPt operates. However, methods such as governmental coordination of all PPE supply, extending or creating new supply through 3D printing all provide viable means of blunting the dearth of PPE in oPt currently.^13, 14^

Our study showed that most HCWs surveyed did not receive adequate training on local protocol or measures to address COVID-19 spread from an institutional perspective. Comparing the preparedness of HCWs in oPt to those around the world, will be a vital element of the debrief from this pandemic and important in developing strategies to ensure the oPt have protocols in place for future public health crises. The lack of current data makes this comparison impossible, currently. In previous pandemics, clinicians in other countries have been substantially more confident in their clinical ability to manage infected patients than what our results reflect; for example, Chinese ICU HCWs during the 2009 H1N1 pandemic were substantially more confident in their preparedness.^15^ This may partly be due to a far greater provision of PPE amongst these workers, that permits greater clinical confidence.

Our study has some important strengths. To our knowledge, this study represents the first attempt to assess the availability of PPE in oPt and the preparedness of HCWs to face the COVID-19 pandemic. We provided a comprehensive evaluation of most PPE described in the literature and used clinically. Participants were well-represented across gender, geographic region, department/specialty, level of training, profession, and type of health care facility.

Potential limitations of this study include small sample size, which may impact generalizability to the greater population of Palestinians. Another weakness of our study was the failure to elicit whether the lack of appropriate PPE was one of the driving factors in reducing HCW confidence in their preparedness. This would then imply attempts to target increasing PPE provision could both protect HCW and improve clinical confidence in managing COVID-19 patients. Potential selection bias arises due to sampling method. Most study participants were recruited from social media posts and emails to the networks of the researchers involved, which may limit some of the study’s generalizability. However, other studies have demonstrated the viability of social media recruitment and snowball sampling to access difficult to reach populations.^16^ Additionally, participants were asked to report on their individual experiences and thus may or may not be wholly representative of the institutions in which they are employed. The cross-sectional nature of this study is also by definition unable to take into account any changes in equipment or training preparedness over time and is only representative of the point-in-time data were collected. These limitations were acknowledged by the authors during study enrolment due to the need to publish findings within the international community in a time-sensitive manner and address the gap in literature regarding COVID-19’s unique impact on the population in the oPt.

## Conclusions

Low-to-middle income countries (LMICs) are particularly vulnerable to the spread of disease because they often grapple with detrimental resource and financial constraints that existed prior to the spread of pandemic. OPt and other LMICs often not only lack proper infrastructure and resources, but also have to navigate restrictions on movement, travel and transportation of essential supplies. In this global pandemic, procurement of adequate supply of PPE and the development of necessary protocols specific to the unique needs and challenges of the region are urgently needed.

## Data Availability

Data are available upon request

## List of abbreviation

COVID-19: Coronavirus disease 19
OPt: Occupied Palestinian territory
HCWs: Healthcare workers
PPE: Personal protective equipment
LMICs: Low-to-middle income countries
WHO: World Health Organization
UNRWA: United Nations Relief and Works Agency
PIEPS: Personnel, Infrastructure, Procedures, Equipment and Supplies
SD: Standard deviation IQR: Interquartile ranges
NGOs: Non-governmental organizations

## Acknowledgement

We would like to thank all participants who took part in this study.

## Declarations

### Ethical approval and consent to participate

Local ethics committee ruled that no formal ethics approval was required. Participants consented to share their responses for research purposes by indicating so when submitting the online survey. Data were kept de-identified to protect participants’ confidentiality.

### Consent for publication

Not applicable.

### Availability of data and materials

The dataset used and analysed during the current study are available from the corresponding author on reasonable request.

### Authors’ contributions

OA contributed to the design of the study, data collection, data analysis, data interpretation and drafting of the manuscript. HA, ZA and AH contributed to data analysis, interpretation and drafting the manuscript. LA and KS contributed to the design of the study and drafting of the manuscript. All authors have read and approved the final manuscript.

### Authors’ information

Ministry of Health, Gaza Strip, occupied Palestinian territory and OxPal Medlink, UK. ^2^Faculty of Medicine, Islamic University of Gaza, occupied Palestinian territory. ^3^Vertex Pharmaceuticals, Boston, USA. ^4^Harvard T.H. Chan School of Public Health, Boston, USA. ^5^Institute for Evidence-Based Healthcare, Faculty of Health Sciences & Medicine, Bond University, Australia. ^6^Medical Sciences Division, University of Oxford and OxPal Medlink, UK.

